# Influence of past infection with SARS-CoV-2 on the response to the BioTech/Pfizer BNT162b2 mRNA vaccine in health care workers: kinetics and durability of the humoral response

**DOI:** 10.1101/2021.05.25.21257788

**Authors:** Jesús Ontañón, Joaquín Blas, Carlos de Cabo, Celia Santos, Elena Ruiz-Escribano, Antonio García, Luis Marín, Lourdes Sáez, José Luis Beato, Ramón Rada, Laura Navarro, Caridad Sainz de Baranda, Javier Solera

## Abstract

**Background:** Vaccines against SARS-CoV-2 have provided an invaluable resource in the fight against this infection. Given the current vaccine shortage, vaccination programs must prioritize their distribution to the most appropriate segments of the population.

**Methods:** We carried out a prospective cohort study with 63 health care workers (HCWs) from a public General Hospital. We compared antibody responses to two doses of BNT162b2 mRNA Covid-19 vaccine between HCWs with laboratory-confirmed SARS-CoV-2 infection before vaccination (experienced HCWs) and HCWs who were not previously infected (naïve HCWs).

**Results:** Seven days after the first vaccine dose HCWs with previous infection experienced a 126 fold increase in antibody levels (GMC 26955 AU; 95% CI 18785-35125). However, in the HCW naïve group the response was much lower and only 5 of them showed positive antibody levels (>50 AU). The HCWs with previous infection did not significantly increased their antibody levels after the second dose while there was a significant increase in the naive HCW group (16 fold; GMC 20227 AU; 95% CI: 15179-25275). Approximately two months after completing vaccination, the level of antibodies was much lower in naive HCWs (GMC 6595 AU vs. 25003 AU; p<0.001)

**Conclusion:** The study shows that 10 months after the disease has passed, the immune system is capable of producing a rapid and powerful secondary antibody response after one single dose of the vaccine. This response reflects the persistence of immunological memory and it is independent of whether or not anti-SARS-CoV-2 antibodies are detected in blood. Besides, we found that the second dose does not improve the antibody response in individuals with previous Covid-19 infection. Nonetheless, two months later, the persistence of antibody levels is still higher in the experienced HCWs. These data suggest that immune memory remains for a long time in recovered individuals, and therefore, vaccination in this group could be postponed until immunization of the rest of the population is complete.

## Introduction

The Covid-19 mRNA vaccine BNT162b2 has shown up to 95% protection against the disease, although the efficacy study excluded participants with a prior clinical or microbiological diagnosis of Covid-19 disease^1^ Current recommendations, however, include individuals recovered from Covid-19 as candidates for vaccination, despite the low rate of reinfection in these individuals shown in cohort studies^2,3^.

An urgent challenge is optimizing vaccine allocation in order to maximize public health benefits. We intend to show that individuals with previous SARS-CoV-2 infection should not be a priority group to receive the vaccine, or, alternatively, it would be sufficient for them to receive one single dose. To achieve this objective, we measured serum anti-SARS-CoV-2 antibody levels as a marker of immune response and we compared the antibody levels in HCW with and without a history of Covid-19 infection. Samples were taken every seven days for three weeks, after the first and second doses of the BNT162b2 vaccine to monitor the immediate and delayed reaction to vaccine administration Serum samples were obtained again two months after the second dose of the vaccine to assess the persistence of anti-SARS-CoV-2 antibody levels

## Methods

The Albacete General Hospital (CHUA) is a 675-bed hospital located in southeastern Spain with 3096 workers. In January 2021, the BNT162b2 vaccine was offered to all of them without any prior exclusion and more than 90% of the personnel received the vaccine according to the standard protocol established by the manufacturer (Pfizer). In an observational prospective longitudinal cohort study, 66 health-care workers (HCWs) agreed to measure their antibody levels before the onset of vaccination and after each of the two doses. The study was approved by the local Clinical Ethics Committee (Internal code: 2021-12 EOm). Informed consent was obtained from all volunteers before sampling. Participants were classified into two groups: “experienced” which included those previously infected with SARS-CoV-2. The date of diagnosis is that of the first positive PCR or the approximate date on which they presented any symptoms compatible with covid-19 prior to the detection of anti-SARS-CoV-2 IgG. None of the individuals in the “naïve” group had clinical or laboratory data suggestive of previous infection. Both groups of participants received the messenger RNA vaccine BNT162b2 (Pfizer–BioNTech).

IgG antibody levels against the SARS-CoV-2 spike region that binds to the receptor-binding domain (RBD) was measured using the SARS-CoV-2 IgG II Quant immunoassay in the ARCHITECT i-System (Abbott, Abbott Park, US). The analytical measurement range is from 21.0 to 80,000 AU / mL and we used the manufacturer’s recommended cutoff point of 50 AU/mL for determining positivity.

To evaluate the kinetics of the immune response, antibody levels were measured at seven time points: a) before or one day after dose one, and at 7, 14 and 21 days after dose one, and b) at 7, 14, and 21 days after dose two. Moreover, to evaluate the persistence of the antibody response, anti-SARS-CoV-2 antibodies were measured again 8 to 10 weeks after the second dose of the vaccine.

Antibody levels were reported using geometric mean concentrations (GMC) with 95% confidence intervals (95% CI). Differences between IgG levels were evaluated using the two-tailed Mann Witney U-test.

## Results

66 HCWs were tested for IgG levels at baseline, before vaccination. Of these, three participants were excluded; one, because she was taking methotrexate and corticosteroids as a treatment for autoimmune disease; and the other one due to rituximab treatment for lymphoma. A third participant was excluded for presenting symptomatic Covid-19 (PCR positive) in the first week after the first dose of the vaccine.

Demographic characteristics of the 63 remaining participants are summarized in Table 1. 33 HCWs had previous Covid-19, 32 of them between March and April 2020, and only one of them had it in September 2020. 5 were asymptomatic, 22 with mild symptoms and another 6 with pneumonia, of which only one of them required oxygen therapy and hospitalization. The median time elapsed between the diagnosis of Covid-19 and the date of the first vaccine dose was 303 days (range 131-338). HCWs with prior Covid-19 were older than HCWs without prior infection. The distribution by sex was the same in the two groups.

**Table 1.**
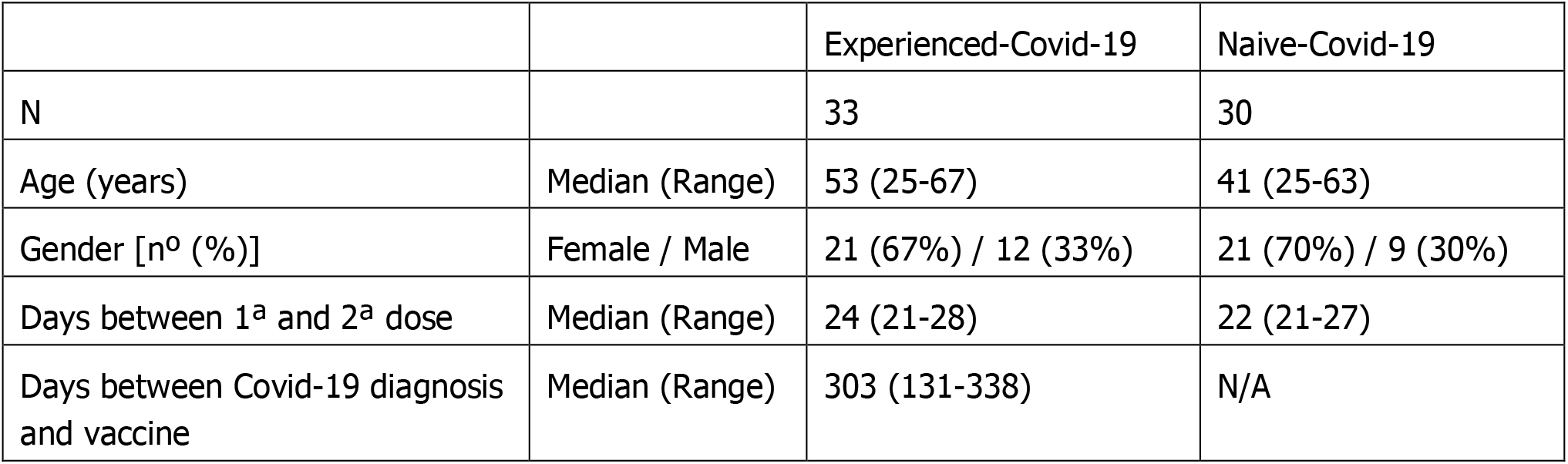
Demographic characteristics of the participants in the study

The results of the measurement of anti-SARS-CoV-2 antibodies are represented in Figure 1. Out of 33 HCWs with previous Covid-19, 29 (88%) had positive (>50 AU) anti-SARS-CoV-2 antibody levels (GMC=311 AU; 95% CI: 144-479). Seven days after they received their first vaccine dose, all of them, including the four seronegative HCWs, experienced an 126 fold increase in antibody levels (GMC 26955 AU; 95% CI 18785-35125; p <0.0001). At day 14 post vaccination, they showed a small, non-significant increase in the anti-SARS-CoV-2 antibody titer (GMC 40701 AU; 95%, CI 34161-47241; p = 0.086) and no increase was observed on day 21. Regarding the HCW “naïve” group, all of them were seronegative in the days prior to vaccination. Seven days after the administration of the first dose of the vaccine, only five of them showed positive IgG values (> 50 AU). At day 14, all of them presented detectable values, although the values reached were still modest (GMC 774 AU; CI: 416-1132; p<0.001). Their levels continued to increase (1.6 fold) until day 21 (GMC 1233 AU; 95% CI 865-1602; p =0.018).

**Figure 1.**
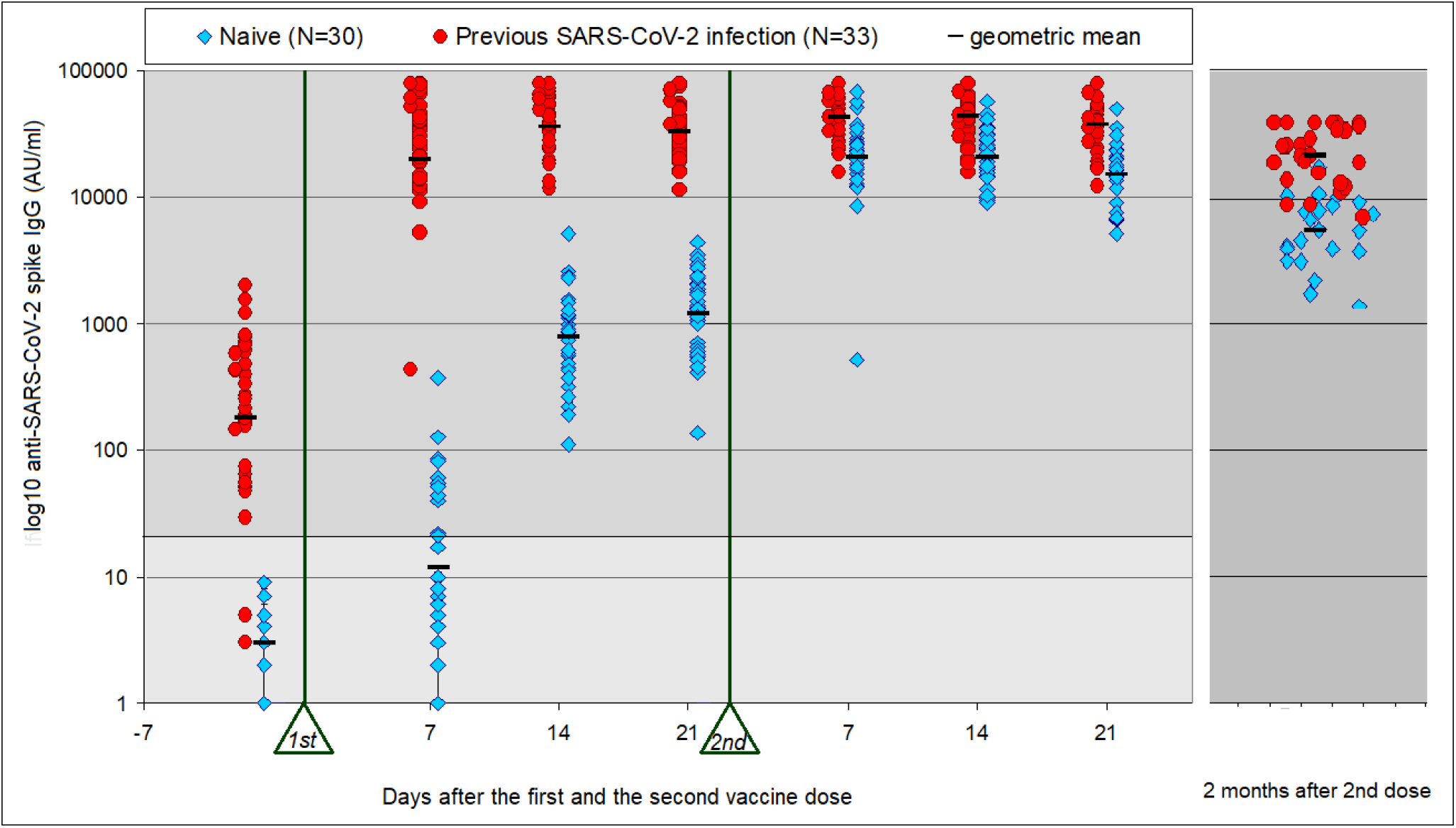
Immunogenicity of the BNT162b2 vaccine (Pfize-BionTech). SARS-CoV-2 (IgG II Quant, Abbott) peak antibody titers from 63 individuals. The moment of administration of the two doses is marked with a triangle and the sampling times are counting from each one of them. The time elapsed between the two vaccine administrations ranged between 21 and 28 days. Some of the individuals with pre-existing immunity had antibody titers below the detection level (21 AU) or below the level established by the manufacturer (50 AU) in the days prior to vaccination.

With regard to the second vaccine administration, there were no significant changes in the antibody levels in the experienced HCW group (Fig1). However, in the group without previous infection another increment was observed, with a 16 fold increase in the antibody titer (GMC 20227 AU; 95% CI: 2576:25275; p<0.001) reflecting a classic secondary response. Also in this group, the titer of antibodies remained stable until day 14 post-vaccination (GMC 20751 AU; 95% CI 16641-24861; NS) but decreased when measured on day 21 post-vaccination (GMC=15408 AU; 95% CI 118016-19001; p=0.036). The antibody titers of HCWs with pre-existing immunity were higher than those in the naïve HCWs at all time points after each dose (p<0.01). The greatest difference was found in the measurements made 7 days after the first dose (2300 fold) and it was gradually reduced to a factor of up to 2 fold by days 14 to 21 after the second dose (Fig. 1)

Within each group, there were no significant differences in the antibody response to the vaccine with regard to sex or age. Neither were there any differences within the group of patients with previous infection between the HCWs who had positive serology before the vaccine and the HCWs who were already negative.

Four HCWs with previous Covid-19 did not receive the second dose of the vaccine; however, they maintained anti-SARS-CoV-2 antibody levels similar to HCWs who were treated with two doses. The data of these 4 HCWs are represented in figure 1, slightly shifted to the left.

Approximately two months (median 66 days; interval 57-86) after the second dose of the vaccine, a new sample was taken for antibody determination and a notable decrease was observed in all cases (Fig. 1). The level of antibodies was much lower in patients without previous infection (GMC 6595 AU vs. 25003 AU; p<0.001). In addition, we calculated the percentage of antibody level decrease for each HCW in relation to their highest titer, finding that the decrease was more pronounced in the HCW naïve group than in the HCW group with previous infection (average 76.46% vs 56%; p<0.0001).

### Reactogenicity to the vaccine

We also compared the frequency of adverse reactions after the first and second vaccination doses in both groups (Fig. 2). The HCWs with previous Covid-19 showed more side effects after the first vaccine administration than the naïve group. However, after the second dose of the vaccine there were no significant differences between both groups. The vaccine was generally well tolerated and medical attention was not required in any case.

**Figure 2.**
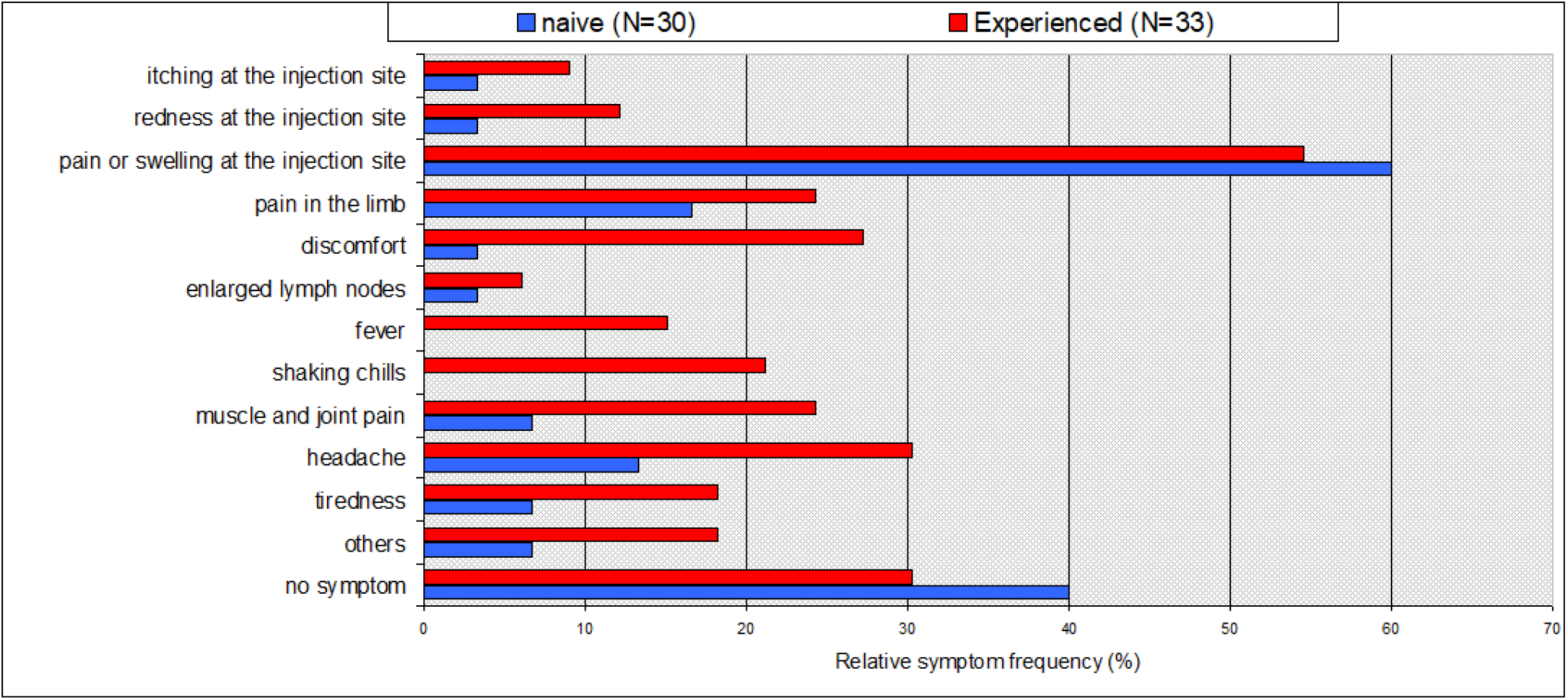
Vaccine-associated side effects experienced after the first dose (N = 66 individuals). Side effects occurred more frequently in people with pre-existing immunity, particularly systemic symptoms. Fever or chills were not observed among patients without pre-existing immunity.

## Discussion

Our study provides evidence for an early, stronger and more durable immune response in Covid-19 experienced HCWs vs. naïve HCWs after vaccination with the BNT162b2 mRNA vaccine. An early and strong response of a similar order of magnitude has been described after the first dose of the vaccine in previous studies in experienced individuals^4,5,6,7,8,9^. It is important to note that this early and strong response was also shown by experienced HCWs who were negative for anti-SARS-CoV-2 antibodies prior to receiving the first dose of the BNT162b2 vaccine. As in previous studies^8,9^, we found that the second dose of the vaccine did not increase anti-SARS-CoV-2 antibody levels in experienced individuals, contrasting with the marked antibody increase still shown by the naïve individuals. Therefore, a single dose may be sufficient for experienced individuals to achieve antibody levels equal to or higher than those obtained by the complete vaccination schedule in individuals without previous infection.

The anti-SARS-CoV-2 antibody levels two months after the end of the vaccination protocol were higher in experienced HCWs vs. naïve HCWs. Moreover, the decline in antibody levels with respect to their peak level was nearly two fold more pronounced in the naïve HCW group. These findings should affect the regimen of administration for potential further doses of the vaccine.

Key limitations of these data are: short follow up time, lack of a recognized correlate of protection, small sample size, none of the participants were older than 67 years old and none of them belonged to racial or ethnic minorities.

Waning antibody levels have been shown in this and recent studies employing anti-SARS-CoV-2 vaccines^10,11^ Longer term data collection is necessary to properly assess this phenomenon. We are currently starting gathering data at regular intervals from individuals up until one year post-vaccination.

In conclusion, our study shows that 10 months after the disease has passed, the immune system is capable of producing a rapid and powerful secondary antibody response after one single dose of the vaccine. Besides, we found that the second dose does not improve the antibody response in individuals with previous Covid-19 infection. Nonetheless, two months later, the persistence of antibody levels is still higher in the experienced HCWs. These data suggest that immune memory remains for a long time in recovered individuals, and therefore, vaccination in this group could be postponed until immunization of the rest of the population is complete.

## Data Availability

Data will be available upon request to the corresponding authors

## Author contributions

Conceptualization: JO, JS, JB, Formal analysis: JO, JS, JB, Investigation: JO, JS, JB, CdC, CS, ER-E, AG, LM, LS, JLB, LN, CSdB, Writing – original draft preparation: CdC, JO, JS, Writing – review and editing: CdC, JO, JS, JB, LM, RR, LS, CS Supervision: JO, CdC, JS

## Acknowledgments

We acknowledge the participation of Álvarez-Gonzalez, María Dolores; Andrés, Carolina; Aretio-Aurtena, Beatriz; Aretio-Aurtenia, Antonio; Arias Martínez, Julio Gabriel; Avilés, María Cortes; Barba, Miguel Ángel; Barrios, María; Benítez, Ramona; Bernabé, Soledad; Blanch, José Javier; Blázquez, José Antonio; Caniego, María Antonia; Carranza, Rafael; Cebrián, Sonia; Collado, María Ángeles; Cortés, José Luís; Dasilva, Beira; Fernández, Daniela; Flor, Ana; García-García, Antonio; García-Sánchez, Manuela; Garbí, Rocío; Gil, Jesús Javier; Gómez-Garrido, Joaquín; González Mozo, Marta; Granero, María Teresa; Guzmán, Marta; Hinarejos, Esther; Jiménez-Hernández, Pedro Antonio; Jiménez-López, Jesús; López-Sanz, Pablo; Lozano, Julia; Martínez-Peral, María Dolores; Mateo, Ángela; Melero, María; Montoya, Julio; Morcillo, María Inmcaculada; Munera, María; Navarro, Amalia; Ochoa Serrano, Alba; Olivas, Otilia; Olucha, Jordi; Parra Ramos, Virginia; Peláez, Celia; Pérez-Trujillo, Andrea; Picazo Montes, María Teresa; Quintanilla, María Pilar; Requena, María Ángeles; Robles, Purificación; Rodríguez-Sánchez, Gemma; Rosa, Cristina; Ruiz-Sánchez, María Pilar; Ruiz-Escribano, Elena; Sáez, Lourdes; Sainz de Baranda, Caridad; Sánchez-Linde, Sergio; Sánchez-Ortiz, Isabel; Segura, Juan Carlos; Solís, Julián Eloy.

The authors also thank the CEO and the Board of Directors of the Albacete General Hospital for their continued support and Alexandra Salewski MSc. for expert English review of the manuscript.

